# Sildenafil in Emergency Treatment of Biliary Colic: A pilot randomized controlled trial

**DOI:** 10.1101/2022.07.27.22277964

**Authors:** Mostafa Yakoot

## Abstract

**Background:** Sildenafil was reported to have a strong inhibitory effect on both gallbladder contraction and biliary pressure of the Sphincter of Oddi. We hypothesized that a single oral dose of sildenafil might counteract the smooth muscle spasm and decrease the intra-ductal and sphincteric pressures to relieve pain of biliary colic and facilitate release of impacted stones.

**Objectives:** To assess the pain-relieving effect of 25 mg oral Sildenafil dose in comparison to an oral 20 mg ketorolac dose in adult patients presenting with acute biliary colic.

**Methods:** Twenty consecutive eligible patients presenting with moderate to severe biliary colic were randomly assigned to receive one Sildenafil 25 mg tablet or two ketorolac 10 mg oral tablets. A four point’s verbal rating scale (VRS) and 100 mm Visual pain analog scale (VPAS) of pain severity were measured before treatment then at 30 and 60 minutes after the dose intake.

**Results:** Treatment success, defined as reduction of VRS, 60 minutes after dose intake from moderate and severe to mild or none was similar (9/10) in both treatment groups. Significant reductions of VPAS at 30 and 60 minutes were noted in both groups. The reduction in VPAS after 60 minutes from dose intake was significantly greater in Sildenafil group (83.16%) than ketorolac group (79.34%) (p=0.033).

**Conclusions:** Single oral dose of sildenafil 25 mg might be safe and effective for relief of biliary colic. Further studies are needed to confirm its value particularly for patients who cannot tolerate or show inadequate response to NSAIDs or opiates.

**Highlights:**

- Sildenafil has a strong relaxing effect on the smooth muscles of gallbladder.
- We tested the effect of sildenafil 25 mg oral dose on the relief biliary colic.
- We found it at least comparable to 20 mg of the potent analgesic ketorolac.
- We recommend further studies on impacted stones in the common bile duct.

## 1 INTRODUCTION

Biliary colic refers to the gallbladder pain that is often located in the epigastrium or right upper quadrant of the abdomen. It is a common cause of presentation to the Emergency Department (ED). It is typically colicky in nature due to muscular spasm of the gallbladder wall secondary to outflow tract obstruction.^**1,2**^

The current acute management options to relieve the severe painful attacks of biliary colic include nonsteroidal anti-inflammatory drugs (NSAIDs) such as ketorolac or opioids such as morphine and meperidine. Both treatment options can provide pain relief in acute biliary colic but not without potential drawbacks. The sphincter of Oddi had been shown to be sensitive to all narcotics that can cause spasm and increase in its pressure. This had been not only demonstrated with morphine but also with meperidine.^**3,4**^

Non-steroidal anti-inflammatory drugs (NSAIDs) including ketorolac are cyclooxygenase enzyme inhibitors that act on prostaglandin release to relieve pain and inflammation. Ketorolac is a potent analgesic non-steroidal anti-inflammatory drug that had been shown to be effective in the treatment of biliary colic ^**5-8**^. These are often administered parenterally in the emergency room, but patients usually prefer to have an oral treatment that can be effective and more convenient.

Sildenafil citrate is a well-known drug used effectively by oral route for the treatment of erectile dysfunction. Sildenafil causes smooth muscle relaxation via increased levels of cyclic guanosine monophosphate (cGMP) by selectively inhibiting “Phosphodiesterase-type 5 isoenzyme (PDE5)”.^**9**^

Sildenafil was proved to have a strong inhibitory effect on contraction of visceral smooth muscle of the gallbladder in healthy volunteers.^**10**^ Sildenafil has also been demonstrated to significantly decrease the basal biliary pressure of the Sphincter of Oddi by more than 60% in individuals undergoing manometry for the evaluation of Sphincter of Oddi dysfunction.^**11**^ Vardenafil, which is another phosphodiesterase type 5 inhibitor acting in the same mechanism as Sildenafil has also demonstrated similar inhibitory effect on the sphincter of Oddi with a significant reduction of the mean basal sphincter pressure and the mean phasic amplitude by more than a half. ^**12**^ We hypothesized that giving a single dose of sildenafil by the preferred convenient oral route during the acute attack of biliary colic, might counteract the smooth muscle spasm which is the main mechanism of biliary colic and probably might decrease the pressure inside the ducts and sphincter of Oddi to relieve pain and may facilitate release of impacted stones.

We conducted a small pilot **hypothesis-generating** clinical study to assess the pain relieving effect of a single-oral dose of 25 mg oral Sildenafil tablet in comparison with a single oral 20 mg ketorolac dose in adult patients presenting with acute biliary colic.

## 2 Patients and Methods

### 2.1 Study design and setting

A randomized, single-blind, active controlled comparative study design, conducted in an outpatient setting.

### 2.2 Patients

A total of 20 eligible consecutive patients presenting with acute biliary colic to the emergency department. The study protocol had been approved by our local institutional review board (IRB00008268) according to the Declaration of Helsinki ethical principles for medical research. All subjects gave written informed consent before performing any treatment intervention. The study was registered with a Clinical Trial Registration ID: ACTRN12619001388101.

Patients were eligible for inclusion if they were adult of any sex, aged between 18 and 50 years old experiencing self-described moderate to severe pain in the epigastrium or right upper quadrant of the abdomen that the treating emergency physician suspected to be biliary colic, plus evidence of gallstones, including a known case record of cholelithiasis or immediate abdominal ultrasound examination showing visible stone(s).

Exclusion criteria included: pregnancy or breast-feeding; history of coronary artery disease or other cardiac disease with hemodynamic instability or fragility; history of intake of any preparation containing nitrates in any dosage form within the past one week; history of allergy to ketorolac, acetylsalicylic acid (ASA) or another NSAID; and history of peptic ulcer disease, gastrointestinal bleeding, perforation, or inflammatory bowel disease; history or evidence of renal or hepatic dysfunctions.

### 2.3 Interventions

All eligible patients were subjected to an initial rapid history taking, physical examination and abdominal ultrasonography to confirm eligibility for inclusion and to collect the pre-treatment demographics, clinical data and pain severity scores. Consenting patients were randomly assigned to receive one Sildenafil 25 mg tablet (experimental group) or two ketorolac 10 mg oral tablets (control group). The randomization scheme was computer generated stratified block randomization by drug and gender and the allocation sequence was concealed by opaque envelopes method. Tablets were handed unlabeled to the patients so only patients were blinded for the type of treatment (single-blind).

Patients were permitted to request rescue medication or drop out of the study at any time if pain relief was inadequate. The study period ended at 60 minutes and a follow up visit or telephone contact was done after 2 days.

### 2.4 Efficacy, safety and tolerability assessment

At 30 and 60 minute intervals after the dose administration of the study medications, the following data were collected by the same attending physician:

Categorical pain assessment: Subjects rated their ongoing pain on a 4 points verbal rating scale (VRS) as (none, mild, moderate or severe).

Visual pain analog scale (VPAS) of pain severity: Subjects marked their pain intensity on a non-demarcated 100 mm horizontal line that had “no pain” written to the left and “most severe pain imaginable” to the right.

Adverse events: Patients were asked to report any adverse events and were asked specifically to grade nausea, vomiting, drowsiness, headache, dizziness, blurring of vision, palpitations, agitation, skin rash, itching and sweating on a 4-point categorical scale. Physiologic parameters: Blood pressure, pulse and respiratory rate were monitored throughout.

### 2.5 End points

The primary end point was successful treatment, defined as the proportion of patients with a change in the VRS from severe or moderate to mild or none at 60 minutes after the dose.

Patients who withdrew from the study or requested rescue medication before the end of the study period were considered treatment failures.

Secondary end points were percent reduction of VPAS scores at 30 and 60 minutes after the dose.

### 2.6 Statistical analysis

Data were analyzed using the computer software package SPSS Statistics for Windows, Version 21.0 (IBM Corporation, Armonk, NY, USA). We presented our qualitative data as counts, proportions or percentage with the confidence interval using Wilson Score Interval Method. For quantitative data, descriptive statistics are the arithmetic mean, the standard deviation, the median and the 95% confidence interval whenever found appropriate.

Because of the small sample size, we used Exact test to test significance of difference between success rates in the 2 treatment groups. Bootstrap for independent samples test was used to compare the percent reduction of VPAS at 30 and 60 minutes in both groups. This work has been reported in line with Consolidated Standards of Reporting Trials (CONSORT) Guidelines.

## 3 Results

Between December 2019 and June 2020, we included 20 eligible patients to the study. All received the study drugs according to the randomization scheme and were closely observed for the full 60 minutes study protocol.

### Baseline characteristics

Baseline demographic and clinical data are summarized in Table 1. The characteristics of the patients in the 2 groups were similar.

**Table 1.** Baseline Characteristics in Both Groups.

|  | **Sildenafil** | **Ketorolac** |
| --- | --- | --- |
| **Randomized (n)** | **10** | **10** |
| **Completed Protocol (n)** | **10** | **10** |
| **Male/female: (n1/n2)** | **3/7** | **3/7** |
| **Age: Mean (SD)** | **35.8 (8.08)** | **33.1 (8.10)** |
| **Body mass index Kg/m<sup>2</sup>: Mean (SD)** | **31.74 (4.7)** | **33.26 (4.4)** |
| **VPAS before treatment: Mean (SD)** | **88.2 (5.09)** | **90.4 (4.06)** |

Abbreviations: n: number; SD: Standard Deviation; VPAS: Visual pain analog score

Successful treatment (VRS pain relief from severe or moderate to mild or none) at 60 minutes occurred in 9 out of 10 patients (90% with 95% confidence interval: 59.6 - 98.2%) in both treatment groups. The difference was not statistically significant (p value = 1.00) by Exact test.

We used the Bootstrap for independent samples test to test the difference in the percent reduction of Visual pain analog score (VPAS) between the 2 treatments after 30 and 60 minutes of dose administration.

After 30 minutes the median percent reduction of VPAS was 56.34% in Sildenafil treated group and 50.1% in Ketorolac treated group. The difference was not statistically significant by Bootstrap independent samples test (P = 0.606) (table 2, 3 and figure 1).

**Table 2.** Percent reduction in VPAS at 30 and 60 minutes in both groups.

| **Group** | **% reduction of VPAS** | **N** | **Mean** | **SD** | **Quartiles** |  |  |
| --- | --- | --- | --- | --- | --- | --- | --- |
|  |  |  |  |  | **Q1** | **Median** | **Q3** |
| Sildenafil | %reduction at 30 m | 10 | 55.58% | 9.71% | 48.69% | 56.34% | 62.22% |
|  | %reduction at 60 m | 10 | 83.16% | 3.42% | 80.91% | 84.09% | 85.18% |
| Ketorolac | %reduction at 30 m | 10 | 53.30% | 9.57% | 45.49% | 50.10% | 61.43% |
|  | %reduction at 60 m | 10 | 78.41% | 5.52% | 76.98% | 78.42% | 81.50% |

Abbreviations: N: number; SD: Standard Deviation; VPAS: Visual pain analog score; m: minutes; Q: quartile

**Table 3.**
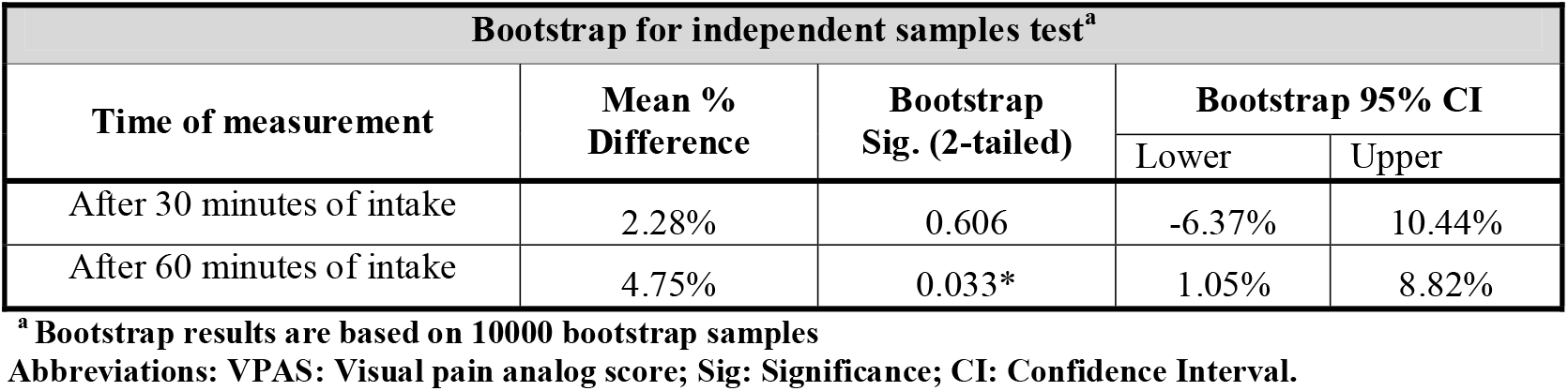
Bootstrap test of Comparison of Percent Reduction of VPAS between groups.

**Figure 1.**
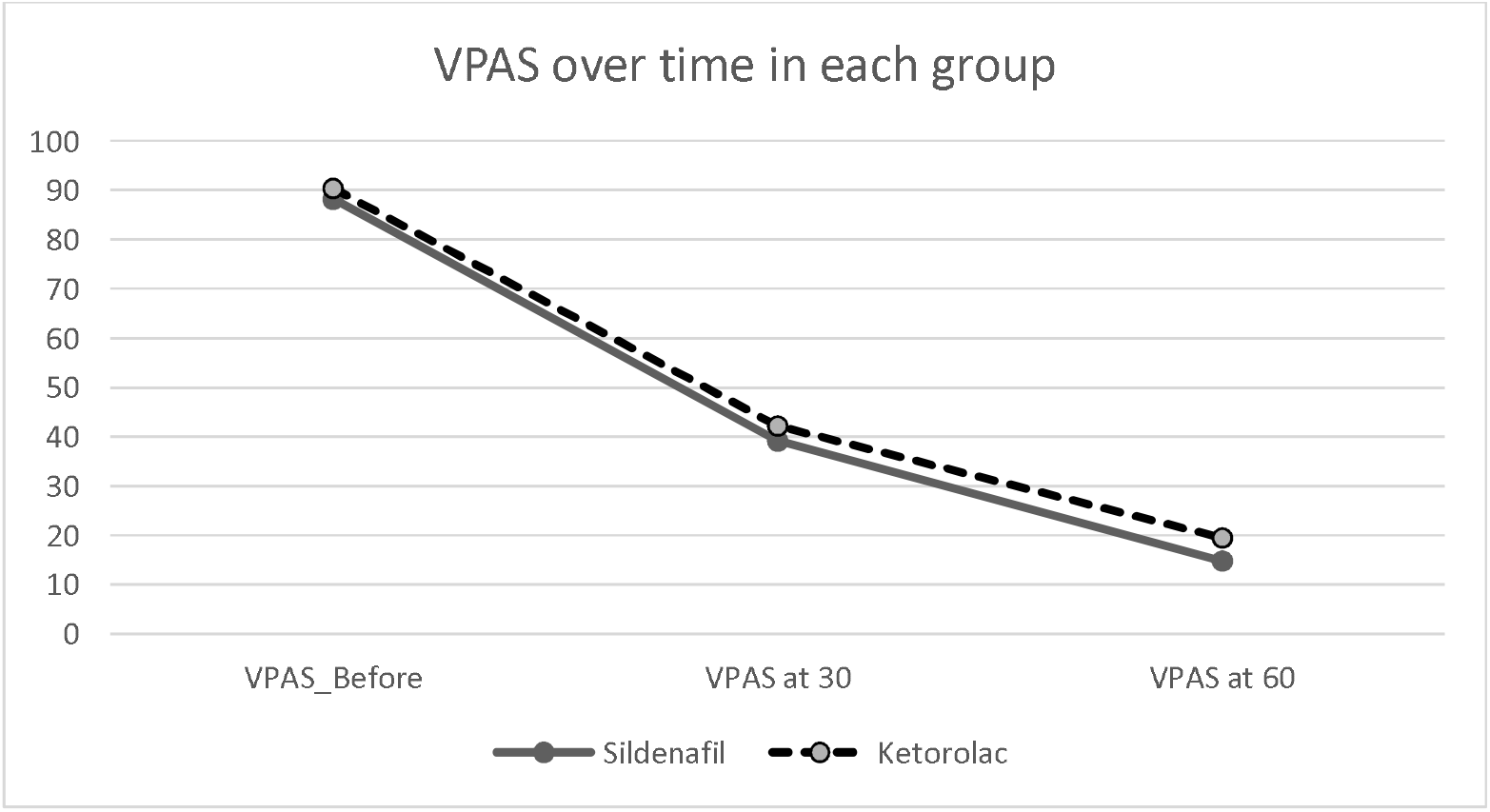
Visual Pain Analog Scores over time in each group

While after 60 minutes of the dose, Sildenafil was associated with a statistically significant (p = 0.033*) greater reduction in VPAS (median = 84.09 %) than Ketorolac (median = 78.42 %).

### Safety and tolerability

Headache, palpitation and flushing were reported by four patients in the Sildenafil treated group versus none in ketorolac group; while feeling of unsteadiness was experienced by 2 in each group. Two complained of nausea and heart burn in sildenafil group versus 3 in ketorolac group. All adverse events were non-serious, short-lived, grade 1 to 2 in severity and all resolved without treatment. We did not receive any further reports of significant adverse events at the Follow up visits or telephone contacts after 2 days of the intervention except 2 patients from ketorolac group and one from sildenafil group complained of recurrence of pain, mild in severity that improved by self-medication with a simple analgesic at home.

## 4 Discussion

In our study the treatment success, defined as reduction of verbal pain scores, 60 minutes after dose administration from moderate and severe to mild or none was 9 out of 10 both treatment groups. The difference was not statistically significant when tested with Exact test based on 10000 iterations (p=1.00).

Both treatment groups showed significant reduction of VPAS 30 minutes after the dose intake. The reduction was slightly greater in the Sildenafil group but the difference was not significant by Bootstrap test for independent samples based on 10000 iterations (p=0.606). The effect at the end of 60 minutes showed a significantly greater reduction in visual pain analog sores in the Sildenafil treated group (83.16%) versus ketorolac group (79.34%). Bootstrap test for independent samples was slightly significant (p=0.033).

We acknowledge the limitation of the small sample size, the short follow up duration and the single-blind design of our study. It has been designed as a small sized pilot hypothesis-generating study that we opted to publish in order to motivate other investigators to replicate it in further larger studies. If the results of this study is further replicated and confirmed by larger studies, these results could allow for the addition of a new option for the emergency treatment of cases suffering moderate to severe biliary colic particularly those who are contraindicated/cannot tolerate; or those who will need a rescue therapy after inadequate response to the use of either NSAIDs or opiates. We also recommend that endoscopists would test the effect of sildenafil (by local or systemic administration) during the endoscopic retrograde cholangiopancreatoscopy (ERCP) management for the impacted common bile duct stones. We expect that it will be a rather non-invasive option in the hands of endoscopists to facilitate the extraction of impacted stones in the common bile duct.

## CONCLUSIONS

Single oral dose of sildenafil 25 mg might be a safe and effective first-aid treatment, for the emergency relief of acute biliary colic in an outpatient setting. Further larger studies are needed to confirm its value as an option added to the armamentarium for the pain relief of moderate to severe biliary colic particularly for patients who are contraindicated/cannot tolerate; or those who will need a rescue therapy after inadequate response to the use of either NSAIDs or opiates.

## Data Availability

All data produced in the present study are available upon reasonable request to the author.

## Notes

**Conflict of interest:** None declared

### Competing Interest Statement

The authors have declared no competing interest.

### Clinical Trial

ACTRN12619001388101 (https://www.anzctr.org.au/Trial/Registration/TrialReview.aspx?id=378014&isReview=true)

### Funding Statement

This study did not receive any funding

### Author Declarations

The study protocol had been approved by the "Green Clinic and Research Center" institutional review board registered by FDA under the number: (IRB00008268) according to the Declaration of Helsinki ethical principles for medical research. All subjects gave written informed consent before performing any treatment intervention.

